# Comparative evaluation of methodologies for estimating the effectiveness of non-pharmaceutical interventions in the context of COVID-19: a simulation study

**DOI:** 10.1101/2024.10.14.24314896

**Authors:** Iris Ganser, Juliette Paireau, David L Buckeridge, Simon Cauchemez, Rodolphe Thiebaut, Mélanie Prague

## Abstract

Numerous studies assessing the effectiveness of non-pharmaceutical interventions (NPIs) against COVID-19 have produced conflicting results, partly due to methodological differences. This study aims to clarify these discrepancies by comparing two frequently used approaches in terms of parameter bias and confidence interval coverage of NPI effectiveness parameters. We compared two-step approaches, where NPI effects are regressed on by-products of a first analysis, such as the effective reproduction number *ℛ*_*t*_, with more integrated models that jointly estimate NPI effects and transmission rates in a single-step approach. We simulated datasets with mechanistic and an agent-based models and analyzed them with both mechanistic models and a two-step regression procedure. In the latter, *ℛ*_*t*_ was estimated first and then used as the outcome in a linear regression with NPI variables as predictors. Mechanistic models consistently outperformed two-step regressions, exhibiting minimal bias (0-5%) and accurate confidence interval coverage. Conversely, the two-step regression showed up to 25% bias, with significantly lower-than-nominal confidence interval coverage, reflecting challenges in uncertainty propagation. We identified additional challenges in the two-step regression method, such high depletion of susceptibles and time lags in observational data. Our findings suggest caution when using two-step regression methods for estimating NPI effectiveness.

## 1 Introduction

The emergence of novel infectious agents, such as the SARS-CoV-2 virus responsible for the COVID-19 pandemic, has highlighted the importance of non-pharmaceutical interventions (NPIs) in mitigating the impact of infectious diseases. NPIs encompass a wide range of public health measures, including social distancing, quarantine, mask-wearing, and school closures, all implemented with the primary goal of reducing disease transmission. The effectiveness of NPIs as a means to mitigate pandemics has been the subject of extensive research during the COVID-19 pandemic.^1–3^ Insights from these studies are crucial in guiding evidence-based public health responses to future pandemics. Various methods and models have been devised to assess NPI impact on disease transmission, ranging from straightforward descriptive techniques^4,5^ and regression models^6,7^ to advanced dynamic models^8,9^ and machine learning approaches.^10,11^ While this diversity of approaches contributes to the robustness of the estimates, it can introduce bias in systematic reviews and meta-analyses if a significant proportion of the methods are potentially unreliable. For example, different estimates of lockdown effectiveness have been found during the first wave in the United States, ranging from no reduction in case growth rates to a reduction by > 50%,^10,12–14^ which can at least partially be attributed to different methodologies being used.

One systematic review reported that the most frequently used methodologies are descriptions of change over time (48% of reviewed studies), non-mechanistic models such as regression models (27%), and mechanistic models (15%). ^15^ Among the latter two, many approaches involve the estimation of intermediary outcomes, primarily the effective reproductive number *ℛ*_*t*_, from raw epidemiological data. These intermediary outcomes are then typically used in regression analyses to derive an estimate of NPI effectiveness. This strategy, which we call “two-step regression approach,”, has been used across a range of studies.^16–20^ Dividing the estimation process into two steps has the advantage of reducing model complexity. However, in addition to the challenges of estimating *ℛ*_*t*_, this approach fails to propagate the uncertainty associated with *ℛ*_*t*_ estimation in the first step to the final estimates. Despite the frequent application of twostep models, the impact of chaining two analysis steps on confidence interval (CI) coverage and parameter bias has not been explored. Moreover, the performance of the one-step approach in estimating NPI effectiveness in mechanistic models remains an open area of investigation, both in terms of parameter bias and correct estimation of uncertainty. ^21^ Here, we describe an extensive methodological study of the two approaches in the context of COVID-19 pandemic inspired by previous results on French data. ^8,19^

## 2 Methods

### 2.1 Study design

Our primary objective was to construct a straightforward example for a meaningful comparison of two methodological approaches. We generated epidemic data both with mechanistic Susceptible-Infected-Recovered (SIR)-type models and agent-based models (ABM) and then compared the performance of mechanistic models with two-step regression models on the simulated data. With each simulation method, we generated a total of 100 datasets, each comprising 94 distinct geographical regions.^8,19^ With both data generation models, we assumed entirely susceptible closed populations. The population sizes for each region were set to the respective population sizes of French departments (range 80k 2.6 million, median 560k). We created scenarios comparable to the first months of an epidemic, with a first NPI, comparable in strength to a lockdown, followed by a second NPI, comparable to a post-lockdown intervention (Figures S4 and S2). Both NPIs were assumed to abruptly reduce transmission on a multiplicative scale, with an immediate and constant effect throughout their implementation.

#### Data generation with a SIR model

We generated data with a SIR model, which consisted of a mathematical model using ordinary differential equations (ODEs) to describe the dynamics of SARS-CoV-2 transmission according to equation 1 and a linear mixed model that determined the transmission rate as a function of NPIs according to equation 2. To allow the basic transmission rate to vary across regions, we included a random intercept.^22^ No measurement error was added to the generated observations. We generated 100 datasets each under five conditions of depletion of susceptibles (2%, 10%, 20%, 40% and 60% depletion of susceptibles before implementation of NPI 1). For parameters used in each scenario, refer to Table S1. The true *ℛ*_*t*_ was calculated as 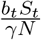, where *b* represents the transmission rate, *γ* the recovery rate, S the number of susceptibles, and N the total population.

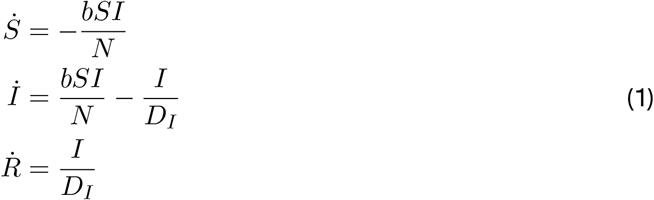

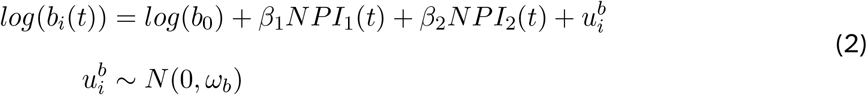

#### Data generation with a SEIRAHD model

To create more realistic scenarios, we generated data with a mechanistic SEIRAHD model, which has been used previously to estimate NPI and vaccine effectiveness.^8,22^ Equation 2 was again used to model the transmission rate as a function of NPIs, and the mathematical model to describe the dynamics of SARS-CoV-2 transmission is presented in equation 3. The mathematical model comprised 7 compartments (Susceptible, latently Exposed, symptomatically Infected, Asymptomatically infected, Hospitalized, Recovered, and Deceased), encompassing various stages of infection (see Figure S1). For a description of the data generation, see Supplementary Methods Section 1.2 and for model parameters, see Table S2. To more closely represent real-life data, we added measurement error to the simulated observations (see Table S3). We kept the initial numbers of infected individuals low in order to have a very low depletion of susceptibles (<2% before implementation of NPI 1).

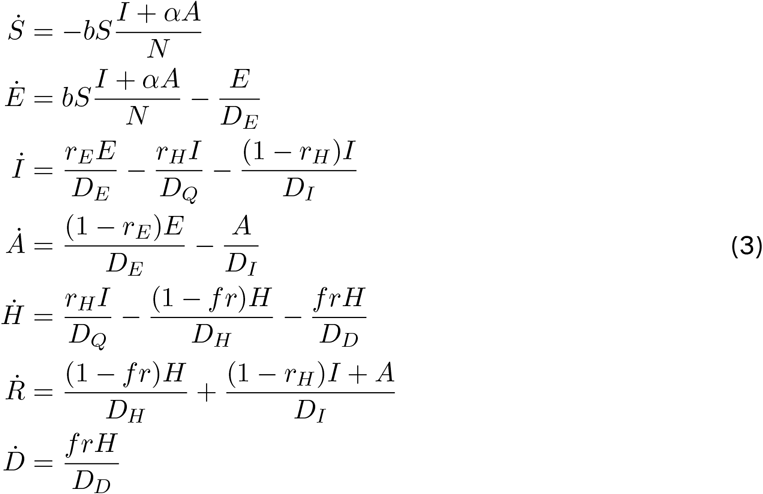

#### Data generation with agent-based model

We generated data with an ABM under two different scenarios: in the random mixing scenario, every agent had an equal probability of coming into contact with any other agent in the population, with an equal probability of transmission for each contact. Conversely, in the multi-layer scenario, interactions were divided into layers of school, workplace, households, and community encounters, with varying transmission probabilities. In the multi-layer scenarios, we assumed that NPIs did not affect household transmission, and disease progression was age-specific. The population size mirrored French departments, and for the multi-layer scenarios, the age distribution and contact structure were set according to the French population. For both scenarios, epidemics were seeded by sampling the number of initially infected agents and the basic viral transmissibility per contact (*vt*) from lognormal distributions (see table S2). Similar to the SEIRAHD models, we kept the depletion of susceptibles very low (2-3%) before the first NPI implementation.

### 2.2 Parameter estimation with mechanistic models

The SIR-generated data were analyzed with the corresponding SIR model. The SEIRAHDgenerated data were analyzed both with the full SEIRAHD model and a reduced SEIR model (described in equation 4).

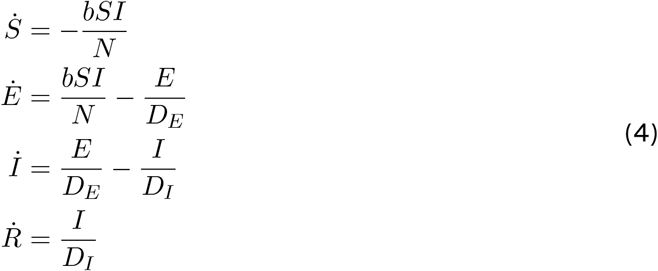

To increase comparability across geographical regions and therefore facilitate estimation, incidence data were scaled to 10,000 population. We fixed the progression parameters in the ODEs (*D*_*I*_, *D*_*E*_, etc.) to their respective true values, while the transmission rate and initial condition of I compartment (SIR model) or E compartment (SEIR/SEIRAHD model) were estimated from the data with random effects, as well as the NPI parameters with fixed effects.

### 2.3 Parameter estimation with two-step regression

The approach for the *ℛ*_*t*_ regression was based on Paireau et al. ^19^ First, we estimated *ℛ*_*t*_ from incident infections or hospital admissions, separately for each simulated region, with a smoothing window of 7 days. In the SIR-generated datasets, we applied no smoothing because the data were generated without measurement error. The approach requires the input of a generation interval. In the SIR model, the generation interval is equal to the *D*_*I*_ parameter, i.e. 5 days. For the data generated with the SEIRAHD model, case and hospitalization data (i.e. entries into the I and H compartments) were used as observations. For both, we calculated a generation interval with a mean of 10.1 days and a standard deviation of 8.75 days according to Wallinga et al. ^23^ (for details, see Supplementary Methods Section 1.3). In the ABMs, we only used symptom onset data for the analysis, and the distribution of the generation interval was calculated directly during simulation, with a mean of 8.45 and a standard deviation of 4.45 for random mixing models and 7.8 and 4.4 for multi-layer models. Second, we ran a mixed-effects regression with the point estimate of the derived log(*ℛ*_*t*_) as outcome and the two NPIs as predictors. Using discretization, for region *i* = 1…94 at weekly time points *j* = 1…17, we modeled:

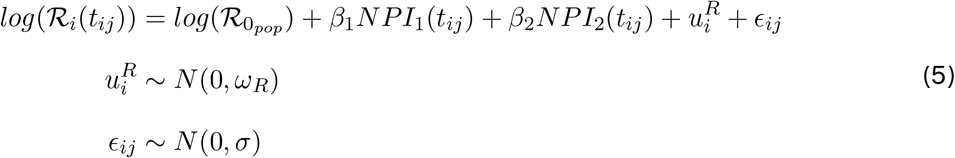

When using data generated with an incubation period (SEIRAHD models and ABMs), we lagged NPIs by 5 days for *ℛ*_*t*_ estimated from cases, and by 10 days for *ℛ*_*t*_ estimated from hospitalizations, to account for transition periods. We performed sensitivity analyses with different lagging periods. We reported the 95% CI using the Normal Distribution, i.e. the mean plus or minus 1.96 times the standard error.

To take into account the uncertainty from the *ℛ*_*t*_ estimation in the regression step, we also implemented a bootstrap procedure by repeatedly sampling from the *ℛ*_*t*_ distribution (details in Supplementary Methods Section 1.4).

### 2.4 Performance evaluation

For comparison of methods, we compared the absolute and relative bias, which we calculated as 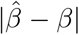 and 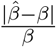, respectively. Additionally, we assessed 95% CI coverage as the percentage of datasets where the 95% CI contained the true value, separately for each estimated NPI parameter.

### 2.5 Implementation

We used the Simulx software version 2021 R2^24^ to simulate the mechanistic model datasets. We used the Python package Covasim version 3.1.4^25^ for ABM simulations, with “new infectious cases” as observations for further analysis. In the mechanistic model approach, parameters were estimated using maximum likelihood estimation using a stochastic approximation expectation maximization (SAEM) algorithm implemented in Monolix. ^24^ Standard errors for calculating 95% CIs were derived from 100 bootstrap samples (by resampling on the 94 geographical regions and varying the algorithm starting point).

The two-step regression analysis was conducted in R version 4.2.3^26^ with the packages EpiEstim^27,28^ using recommendations from references^29^ and^30^ to estimate *ℛ*_*t*_ and lme4^31^ for the mixed effects regression. All code is publicly available on GitHub (https://github.com/sistm).

### 2.6 Bias exploration

To detect possible issues in the regression step, we ran linear mixed models with the true *ℛ*_*t*_ as the outcome variable. In the SEIRAHD-created datsets, the true *ℛ*_*t*_ was calculated as a linear transformation of the transmission parameter, using the next generation matrix approach (see Supplementary Methods Section 1.5). ^32^ In the ABM datasets, *ℛ*_*t*_ was computed directly during the simulation as the quotient of new infections on day *t* over the number of infectious agents on the same day, multiplied by the average duration of infectiousness.^25^

To investigate the potential impact of NPI strength and implementation time on the two-step model performance, we simulated data with diverse NPI implementation times (ranging from a 20-day to a 60-day NPI-free period) and varied NPI 1 strengths (coefficients ranging from −0.5 to −2, corresponding to a percentage reduction in transmission between 39% and 86%).

## 3 Results

### 3.1 Exploring bias in the two-step regression models

#### Data created with SIR model

First, we analyzed data generated with a simple SIR model, and different scenarios of depletion of susceptibles (ranging from 2% to 60%). We found that the bias in NPI effect estimations from the two-step regression model increased with greater depletion of susceptibles, whereas the mechanistic model consistently estimated the correct value (Table 1). For example, with a 2% depletion of susceptibles, the bias of the two-step regression model in estimating NPI 1 was 1%, which increased to 15% at 20% depletion of susceptibles and 45% at 60% depletion of susceptibles. Moreover, the 95% CI of the mechanistic model covered the true value in all 100 simulated datasets. In contrast, the CIs from the two-step regression procedure were consistently too narrow, failing to cover the true value even in scenarios with little bias. The CI width was improved by bootstrapping the two-step regression procedure, but adequate coverage was only achieved in the scenario with the least bias. Of note, in the 40% and 60% depletion of susceptible scenarios, the 95% CIs for NPI 2 showed good coverage despite a large bias. This anomaly can be attributed to the absence of viral transmission during the NPI 2 period, due to the high prior depletion of susceptibles (illustrated in Figure S5). Consequently, NPI 2 could only be estimated with high uncertainty, with 95% CIs ranging from −2.57 to −0.37, corresponding to a percentage reduction in transmission from 31% to 92%, making the CIs so wide that they are practically meaningless (Figure S6).

**Table 1:** Evaluation metrics from SIR simulation. For each scenario of depletion of susceptibles, the mean absolute and relative bias and percentage of CIs covering the true value across 100 simulated datasets are shown. The columns indicate the analysis model. The CI rows show the percentage of datasets where the 95% CI covers the true value. The 95% CI of the mechanistic model was always determined with bootstrap. Reg. two-step regression model, Mech. mechanistic model, CI confidence interval, NPI nonpharmaceutical intervention

| Depletion of S | 2% |  | 10% |  | 20% |  | 40% |  | 60% |  |
| --- | --- | --- | --- | --- | --- | --- | --- | --- | --- | --- |
|  | Reg. | Mech. | Reg. | Mech. | Reg. | Mech. | Reg. | Mech. | Reg. | Mech. |
| **NPI 1** |  |  |  |  |  |  |  |  |  |  |
| Absolute bias | -0.02 | 0.00 | 0.10 | 0 | 0.21 | 0 | 0.40 | 0 | 0.65 | 0 |
| Relative bias (%) | 1.2 | 0.2 | 7.0 | 0 | 14.8 | 0 | 27.4 | 0 | 45.0 | 0 |
| 95% CI (%) | 0 | - | 0 | - | 0 | - | 0 | - | 0 | - |
| 95% bootstrap CI (%) | 100 | 100 | 0 | 100 | 0 | 100 | 0 | 100 | 0 | 100 |
| **NPI 2** |  |  |  |  |  |  |  |  |  |  |
| Absolute bias | 0.05 | 0 | 0.20 | 0 | 0.33 | 0 | 0.42 | 0 | 0.48 | 0 |
| Relative bias (%) | 6.6 | 0.1 | 24.5 | 0 | 40.9 | 0 | 51.9 | 0 | 59.5 | 0 |
| 95% CI (%) | 0 | - | 0 | - | 0 | - | 0 | - | 0 | - |
| 95% bootstrap CI (%) | 100 | 100 | 0 | 100 | 0 | 100 | 100 | 100 | 100 | 100 |

The influence of the depletion of susceptibles on the bias of estimates can be understood analytically. In the two-step regression procedure, NPI effects were estimated using the *ℛ*_*t*_ estimated in the first step according to equation 5. With 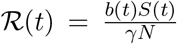 and replacing *b* by equation 2, we derive:

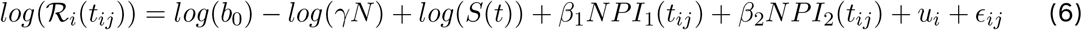

In this equation, *log*(*b*_0_) and *log*(*γN*) are constants and thus included in the intercept term. In contrast, *log*(*S*(*t*)) is time-varying and thus has the potential to bias the estimated NPI effects, with a greater depletion of susceptibles over the estimation period leading to an increased bias.

#### Data created with SEIRAHD model

While the SIR scenarios are useful to understand the general underlying challenges of the two-step regression procedure, the SIR model’s simplicity does not capture the complexity of real-world scenarios. The data generated by the SEIRAHD model address this limitation by offering a more realistic representation of an epidemic. The point estimates from the two-step regression models displayed substantial bias, particularly pronounced for the first NPI (relative bias ranging from 18-25%) compared to the second NPI (approximately 14-18%, see Table 2). Throughout all datasets, using hospitalizations for *ℛ*_*t*_ estimation and subsequent regression consistently resulted in higher bias compared to using case data. Moreover, the CIs derived from these models consistently failed to include the true NPI values. When the two-step regression procedure was bootstrapped, the CIs were wider and included the true value for NPI 2, but not for NPI 1.

**Table 2:** Evaluation metrics over 100 datasets created with the mechanistic SEIRAHD model. The columns indicate the analysis model. The CI rows show the percentage of datasets where the 95% CI covers the true value. The 95% CI of the mechanistic model was always determined with bootstrap. CI confidence interval, hosp. hospitalization, NPI non-pharmaceutical intervention

| Metric | SEIR model | SEIRAH model | Regression model cases | Regression model hosp. |
| --- | --- | --- | --- | --- |
| **NPI 1** |  |  |  |  |
| Absolute bias | 0.00 | 0.01 | -0.26 | -0.37 |
| Relative bias (%) | 0.3 | 0.4 | 18.3 | 25.4 |
| 95% CI (%) | - | - | 0 | 0 |
| 95% bootstrap CI (%) | 100 | 100 | 0 | 0 |
| **NPI 2** |  |  |  |  |
| Absolute bias | 0.01 | 0.00 | -0.11 | -0.15 |
| Relative bias (%) | 0.8 | 0.7 | 13.7 | 18.5 |
| 95% CI (%) | - | - | 0 | 0 |
| 95% bootstrap CI (%) | 100 | 100 | 99 | 100 |

In contrast, the 95% CIs for both NPIs derived with the mechanistic models covered the true value in all 100 datasets, while the point estimates exhibited only minimal absolute and relative bias (<1% for both NPIs, detailed in Table 2). The exceptional accuracy of the SEIRAHD model was anticipated, as it was the model used for data generation.

### 3.2 Origins of bias

In light of the substantial bias observed in the two-step regression model when a more realistic model was used for data generation, we investigated in depth the origins of this issue. Firstly, we examined the regression analysis step by running the linear mixed-effects model with the true *ℛ*_*t*_ values as the outcome variable. While the regression model fitted the true *ℛ*_*t*_ almost perfectly and estimated NPI effects with only slight bias for data generated by the SEIRAHD model (Table S4 and Figure 1A), the CIs failed to cover the true values due to the estimation of extremely small standard errors. However, based on these findings, we ruled out the regression step as the primary contributor to the bias.

**Figure 1:**
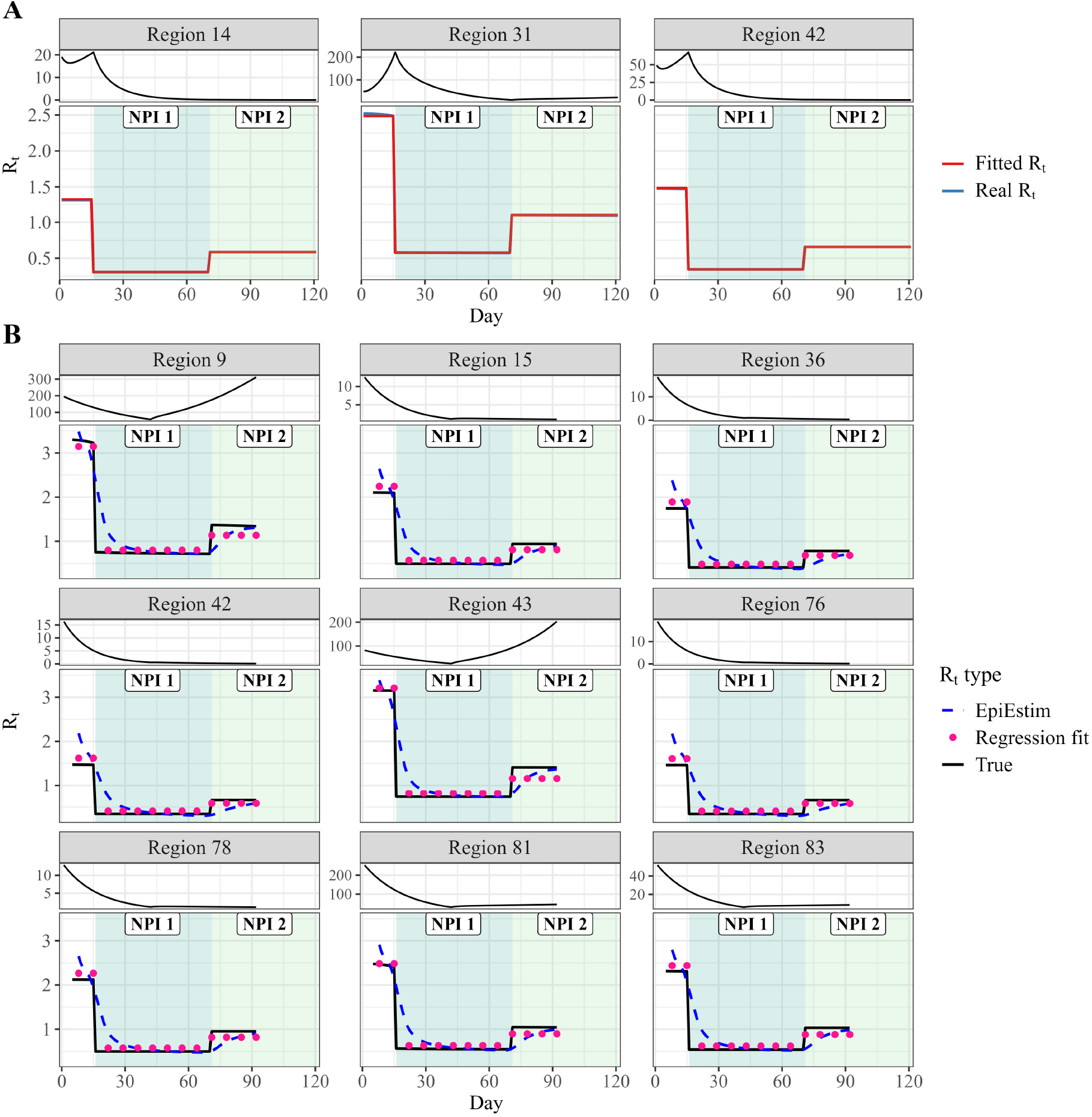
2-step regression bias exploration. **A:** Regression fits of true ℛ_*t*_ in three randomly selected regions. Each panel represents one geographic region with data generated by the mechanistic SEIRAHD model. The true ℛ_*t*_ is depicted in blue and the corresponding regression fit in red. The panels on top show the respective case time series. **B:** ℛ_*t*_ fits by the two-step procedure and subsequent regression for data generated by the mechanistic SEIRAHD model. Each panel represents one geographic region. The highlighted regions indicate which NPI was active at which time. The top panels the respective case time series. Note that we followed EpiEstim guidelines in terms of not estimating ℛ_*t*_ before 2 generation times after the start of the epidemic, but these 2 weeks are cut off from the plot.

Comparing the *ℛ*_*t*_ curves estimated in the two-step procedure to the true *ℛ*_*t*_ from the mechanistic SEIRAHD model, we identified discrepancies at the onset of the epidemic and a lag in *ℛ*_*t*_ estimation by EpiEstim when the true *ℛ*_*t*_ underwent sudden changes resulting from the implementation or lifting of NPIs (Figure 1B). These lags led to an underestimation of the strength of NPI 1 and overestimation of NPI 2, as the regression model estimated an average of the NPI periods. The pronounced decline in the first days contributed to the regression model consistently overestimating *ℛ*_0_, i.e. *ℛ*_*t*_ at the onset of the epidemic.

We proceeded to investigate whether NPI strength had any discernible impact on the bias in *ℛ*_*t*_ estimation. For NPI 1, we observed that both absolute and relative bias increased with the rise in NPI strength. Regarding NPI 2, the bias followed a U-shaped pattern with increasing NPI 1 strength, with an underestimation of *ℛ*_*t*_ during the NPI 2 period by all models (Figures S8 and S9). A more gradual NPI implementation period, involving a linear increase and decrease of NPIs from 0 to 1 over 1 or 2 weeks, did not improve *ℛ*_*t*_ estimation nor the bias in regression coefficients (Figure S10 and Table S5).

Since the SIR model produced accurate results in the scenarios with low depletion of susceptibles, we hypothesized that a potential source of error in the two-step procedure could stem from the convolution of the time series. For an optimal *ℛ*_*t*_ estimation, the most pertinent data are the dates of infection, aligning with the entry into the E compartment in our model, thereby capturing real-time transmission dynamics. Estimating *ℛ*_*t*_ based on newly infected (corresponding to entry into the E compartment) instead of newly symptomatic (entry into I compartment) resulted in a notable reduction in relative bias for NPI 1, diminishing to 4.5%. However, the bias in NPI 2 estimation increased to 22.9% (see Table S6).

### 3.3 Limitations of the mechanistic approach in the context of misspecified models

To assess the robustness of the mechanistic model approach in the face of model misspecification, we generated data with ABMs, which include more heterogeneous individual behavior and population interactions, and a different underlying disease progression than assumed in the SEIRAHD model. We observed that even within the ABM framework, the mechanistic SEIR model in general demonstrated superior performance in terms of bias and coverage compared to the two-step regression model. The SEIR model effectively estimated NPI 1 with minimal bias around 2% and 95% CIs covered the true value in more than 95% of datasets, regardless of whether the data were generated using random mixing or the multi-layer ABM (Table 3). However, for NPI 2, CI estimated by the SEIR model covered the true value in only 71% of the random mixing datasets but 100% of the multi-layer datasets. For NPI 1, the CIs derived from the regression model (both bootstrapped and non-bootstrapped) systematically failed to cover the values and displayed significant underestimation (relative bias of 12% for random mixing and 19% for multi-layer). However, the bias for NPI 2 was substantially lower (5% for random mixing and 1% for multi-layer).

**Table 3:** Evaluation metrics for 100 datasets created with the agent-based model. The CI rows show the percentage of datasets where the 95% CI covers the true value. The 95% CI of the mechanistic model was always determined with bootstrap. ABM agent-based model, CI confidence interval, NPI non-pharmaceutical intervention, reg regression

|  | SEIR random mixing | SEIR multi-layer | Reg model random mixing | Reg model multi-layer |
| --- | --- | --- | --- | --- |
| **NPI 1** |  |  |  |  |
| Absolute bias | 0.04 | -0.02 | -0.18 | -0.27 |
| Relative bias (%) | 2.6 | 1.3 | 12.2 | 18.7 |
| 95% CI (%) | - | - | 0 | 0 |
| 95% bootstrap CI (%) | 100 | 100 | 0 | 0 |
| **NPI 2** |  |  |  |  |
| Absolute bias | -0.04 | 0.02 | -0.05 | -0.01 |
| Relative bias (%) | 4.7 | 3.2 | 5.7 | 1.5 |
| 95% CI (%) | - | - | 0 | 91 |
| 95% bootstrap CI (%) | 71 | 100 | 0 | 95 |

## 4 Discussion

We evaluated and contrasted the performance of mechanistic models with two-step *ℛ*_*t*_ estimation and subsequent regression modelling for estimating the relative reduction in viral transmission caused by NPIs. Mechanistic models consistently outperformed the two-step procedure both in terms of bias and coverage. The two-step procedure consistently underestimated standard errors of the parameter estimates across all analyses. This issue stems from the failure to propagate the error in *ℛ*_*t*_ estimation into the final estimate, compounded by the overconfidence of the regression procedure, as observed in regressions with known *ℛ*_*t*_ as the outcome variable. We showed that this issue could be improved by repeatedly sampling from the posterior distribution of the *ℛ*_*t*_ estimated in the two-step procedure.

Similar to Gostic et al., ^30^ we found that in a basic SIR scenario without weekly smoothing of observations and low depletion of susceptibles, *ℛ*_*t*_ was estimated accurately, leading to nearly unbiased NPI effectiveness parameters. This result suggests that the parameter bias observed in the two-step regression model was not uniform across scenarios. However, in scenarios with higher depletion of susceptibles, the bias increased substantially. As an epidemic progresses, the number of susceptibles diminishes, resulting in a reduction of *ℛ*_*t*_. While not problematic for *ℛ*_*t*_ estimation itself, the regression procedure will attribute the decrease in *ℛ*_*t*_ to the NPI, thus making them appear more effective than they truly are, with the bias increasing as the depletion of susceptibles increases.

In the more realistic scenarios, such as those generated by the SEIRAHD and ABM models as compared to the scenarios generated by SIR models, we observed greater bias in the point estimates produced by the two-step regression procedure, particularly for the first NPI. This bias can be attributed to several factors. First, the representation of the natural history of infection in the SEIRAHD model and ABM differs from that assumed by EpiEstim. If we had generated data with a mechanism more consistent with EpiEstim, i.e., with the generation time distribution as an input parameter, estimation with the SEIRAHD model would likely have resulted in bias for the mechanistic models. This is because misspecification of the generation time distribution can bias estimates of the reproduction number, regardless of the approach used. ^23^ However, it remains debatable which approach is more realistic: simulating with the generation time as an input parameter or simulating with an underlying compartmental structure.

Second, the inability to replicate the sharp decline induced by NPI implementation can be attributed to the long smoothing time window (7 days) coupled with a lengthy generation interval (10.1 days in SEIRAHD models). This gradual convergence of the estimated to the true *ℛ*_*t*_ following NPI implementation, led to inaccurate estimations of NPI impact, as regression models fit an average across the entire NPI period. However, we found that gradually implementing NPIs did not reduce the bias in regression estimates. Moreover, smoothing is necessary to manage measurements errors and other irregularities in the observational data.

Third, the lag in observational time series behind real-time transmission might contribute to the bias, as symptomatic infections or hospitalizations capture transmission events that occurred in the past. This delay cannot be rectified by merely lagging the NPIs, and could explain why estimates from hospitalizations were less accurate than estimations from cases, as we only shifted NPI periods without considering the deconvolution of the time series.^30^ Indeed, using transmission-related observations directly (entry into the E compartment) helped reduce this bias. Several R packages for back-calculating transmission events from cases or hospitalizations are now available, such as EpiNow2 and EstimateR.^33,34^

Using regression analyses without accounting for the depletion of susceptibles also precludes strong causal conclusions about the effect of NPIs. Mechanistic models, which explicitly consider viral transmission mechanisms and therefore depletion of susceptibles, offer an alternative for causal interpretation, ^21^ but require detailed data and time to develop and estimate models. Running 100 bootstrap repetitions on 100 SIR datasets parallelized on 20 high-performance computing nodes took approximately 42 hours. Since the two methodologies were run on different computing platforms, their computing times are hard to compare. Nevertheless, the two-step regression procedure, parallelized on 16 conventional laptop cores, required only four hours of computing time. In an early epidemic or pandemic setting, timely results are of great importance, so this trade-off between speed and accuracy of the results needs to be taken into account when deciding on a model. Therefore, developing user-friendly software for rapid epidemiological modeling in such scenarios is essential.

Our study comes with limitations that need to be acknowledged. First, it is important to note that our simulations do not prove that the mechanistic approach will always be unbiased. Indeed, in estimating parameters in datasets created by ABMs, we observed a reduced CI coverage with mechanistic models. Second, our simulated datasets did not consider various systematic biases, such as reporting delays, significant under-reporting or missing observations. The only measurement error present was random noise on observations, and we did not incorporate weekly trends or seasonal changes in transmission. Moreover, we simulated only two consecutive NPIs with no overlap. Our most realistic scenarios were therefore simpler than real-life scenarios during the COVID-19 pandemic, with spatial structures, multiple overlapping NPIs implemented to varying degrees, behavioural dynamics, and more. It is likely that in a reallife scenario, the problem could be even more exacerbated because of practical identifiability issues. However, our primary objective was to illustrate and compare the performance of two analysis methods under close-to-optimal conditions, and these limitations to not threaten the validity of our results. To address some of these simplifications, we included simulations using ABM. However, we acknowledge that when analyzing real-world data, misspecification of the mechanistic model (for example, assumptions about the natural history of infection) might equally lead to bias. This is particularly true in the context of real-time modelling of emerging pathogens.

Improving the public health response during an epidemic depends on informed decision-making about NPIs. Our findings have significant implications for refining the methodology used to estimate the effectiveness of NPIs. Our findings highlight the potential for a systematic underestimation of uncertainty in the two-step regression procedure, raising concerns about the reliability of its effectiveness estimates across different scenarios. While compartmental models demonstrate superior performance over simpler models, their resource requirements, as they also require more time and expertise to implement, must be weighed against their benefits.

## Supporting information

All appendix files

## Data Availability

All data produced in the present work are simulated. Data produced in the study are available upon reasonable request to the authors or can be re-simulated from the scripts available on GitHub (https://github.com/sistm/SEIR\_vs\_RTreg)

## 5 Contributions

Conceptualization: MP, RT, IG, SC and supervision: MP, DLB, and RT. Formal analysis, writing - original draft: IG. Methodology and writing - review & editing: all.

## 7 Data sharing

All code is available at the SISTM team’s GitHub (https://github.com/sistm/SEIR_vs_RTreg).

## 8 Acknowledgements

IG is supported by the Digital Public Health Graduate Program within the framework of the PIA3 (Investment for the Future), project reference: 17-EURE-0019, and by a doctoral award from the Fonds de recherche du Québec-Santé. This work has been pursued in the EMERGEN project framework of the French Agency for Research on AIDS and Emerging Infectious Diseases (ANRS0151) and supported by INSERM and the Investissements d’Avenir program, Vaccine Research Institute (VRI), managed by the ANR under reference ANR-10-LABX-77-01. We thank Lixoft SAS for their support. Numerical computations were in part carried out using the PlaFRIM experimental testbed, supported by Inria, CNRS (LABRI and IMB), Université de Bordeaux, Bordeaux INP, and Conseil Régional d’Aquitaine (see https://www.plafrim.fr).

