## Supplementary material for "Comparative evaluation of methodologies for estimating the effectiveness of non-pharmaceutical interventions in the context of COVID-19: a simulation study": All appendix files

#### 1 Supplementary Methods

##### 1.1 SIR models

| Parameter | Interpretation | Scenario | Value |
|---|---|---|---|
| $b_0$ | Basic transmission rate | all | $\sim \mathcal{N}$(0.425, 0.01) |
| $D_I$ | Infectious period (days) | all | 5 days[35] |
| $\gamma$ | recovery rate (1/days) | all | 0.2 (= 1/$D_I$) |
| log(initI) | initial number of infected individuals per region | 2% S depletion | $\sim \mathcal{N}$(-3, 0.4) |
| | | 10% S depletion | $\sim \mathcal{N}$(-1.2, 0.4) |
| | | 20% S depletion | $\sim \mathcal{N}$(-0.4, 0.4) |
| | | 40% S depletion | $\sim \mathcal{N}$(-0.6, 0.4) |
| | | 60% S depletion | $\sim \mathcal{N}$(-1.6, 0.4) |
| $\beta_1$ | Effectiveness parameter of NPI 1 | all | -1.45[8] |
| $\beta_2$ | Effectiveness parameter of NPI 2 | all | -0.8[8] |

Table S1: Parameters governing the SIR model

##### 1.2 SEIRAHD model

The values of parameters governing the SEIRAHD model were sampled from prior distributions to achieve different realizations of epidemics: We seeded the epidemic in each region by randomly sampling an initial number of exposed, and set the initial values of the I, A, H, and R compartments as functions of this number. The D compartment was assumed to be empty at the beginning of simulations, and S was set to complete a population size of the geographical region. From the seeded compartment values and basic transmission rate, our model simulated daily compartment values deterministically, under the assumption of random mixing and uniform disease progression. The values of all model parameters can be found in Table S2.

We assumed full reporting of all infected cases, hospitalizations, and deaths. However, to account for measurement errors in epidemiological data, random noise was added to generated observations. This was achieved by randomly sampling a value from a distribution around each the simulated observation, which used a combined error model of the form $y = x + (a + bx)\epsilon$, where $x$ was the "true" value for each observation, $a$ was a constant error term, and $b$ was a proportional error term, denoting that the error amplitude increases with the predicted value's magnitude (see table S3). Exemplary datasets of the resulting simulations are shown in Figure S2

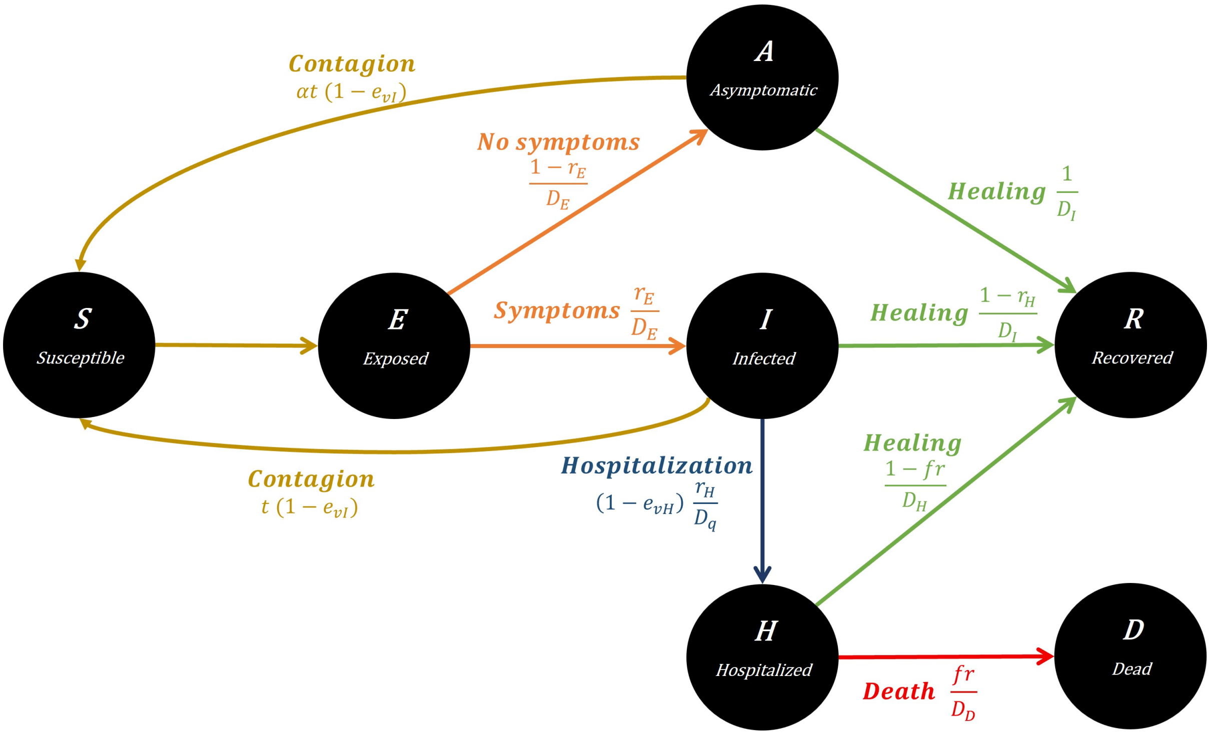


Figure S1: Flowchart of SEIRAHD model

##### 1.3 Generation intervals

In the SEIRAHD model, several distributions of waiting times have to be taken in to account to calculate the distribution of the generation interval. The SEIRAHD model used to generate the data has two infectious compartments (A, I) and two compartments which infectious individuals can progress to (R, H). However, very conveniently, the waiting times in all infectious compartments ($A \rightarrow R$, $I \rightarrow R$, $I \rightarrow H$) are all exponentially distributed with a mean of 5 days (and therefore a rate parameter of 0.2). Therefore, the three above processes can be combined into one, and the SEIRAHD model can be simplified to a SEIR model for the calculation of the generation interval.

Following Wallinga et Lipsitch,[23] there is a relationship between the reproductive number $\mathcal{R}_t$, the growth rate $r$, and the generation interval GI (with its distribution $g(a)$). More specifically, $\mathcal{R}_t$ is related to $r$ according to the moment-generating function (MGF) of the GI $M_{g(a)}$. A MGF, if it exists, uniquely characterizes the shape of the entire probability distribution: $M(z)$ determines $g(a)$ and, conversely, $g(a)$ determines $M(z)$. Thus, in order to find the distribution $g(a)$, we can use the corresponding MGF $M_{g(a)}$.

For an exponentially distributed random variable of mean $1/\lambda$, the moment generating function is $M(z) = \frac{\lambda}{\lambda - z}$. For successive stages of disease (E and I in the SEIR model), the MGFs of

| Parameter | Interpretation | Value |
|---|---|---|
| **Mechanistic Model** | | |
| $r_E$ | Ratio of symptomatic cases among all infected | 0.85 [36] |
| $r_H$ | Hospitalization rate | 0.1 |
| $D_E$ | Latent (incubation) period (days) | 5.1 days [37] |
| $D_I$ | Infectious period (days) | 5 days [35] |
| $\alpha$ | Ratio of transmission between *A* and *I* | 0.55 [38] |
| $D_Q$ | Duration from infection to hospitalization (days) | 5 [8] |
| $D_H$ | Length of stay in hospital (days) | 18 [22] |
| $D_D$ | Duration from hospital admission to death (days) | 10 [39] |
| $fr$ | Death rate of hospitalized patients | 0.1 [8] |
| log(initE) | initial number of exposed individuals per region | $\sim \mathcal{N}$(2, 0.9) [8] |
| $b_0$ | Basic transmission rate | $\sim \mathcal{N}$(0.5, 0.1) [8] |
| $\beta_1$ | Effectiveness parameter of NPI 1 | -1.45 [8] |
| $\beta_2$ | Effectiveness parameter of NPI 2 | -0.8 [8] |
| **ABM** | | |
| vt (random mixing) | Basic viral transmissibility per contact in random mixing models | $\sim log\mathcal{N}$(0.016, 0.02) [25] |
| vt (multi-layer) | Basic viral transmissibility per contact in multi-layer models | $\sim log\mathcal{N}$(0.017, 0.02) [25] |
| Initial exposed | Initial number of exposed individuals per region | $\sim \mathcal{N}$(50, 5) |

Table S2: Parameters governing the more complex data generation models

individual stages can be chained to calculate the generation interval:

$$M_g(z) = M_E(z) \times M_I(z) = \frac{\lambda_E}{\lambda_E - z} \times \frac{\lambda_I}{\lambda_I - z} \tag{1}$$

where $M_E(z)$ is the MGF of the time spent in compartment E and $M_I(z)$ is the MGF of the time spent in compartment I.

The first derivative of the MGF evaluated at z=0 is the mean of the generation interval. In our case:

$$M_g'(0) = \frac{1}{\lambda_E} + \frac{1}{\lambda_I} \tag{2}$$

With $\lambda_E = \frac{1}{5.1} day^{-1}$ and $\lambda_I = \frac{1}{5} day^{-1}$, we thus obtain $E[GI] = 5.1 + 5 = 10.1$.

The second derivative of the MGF evaluated at z=0 is the variance of the generation interval.

$$M_g''(0) = \frac{1}{\lambda_E^2} + \frac{1}{\lambda_E \lambda_I} + \frac{1}{\lambda_I^2} \tag{3}$$

| Parameter | Interpretation | Value |
|---|---|---|
| $a_c$ | additive error cases | 0.0408 |
| $b_c$ | multiplicative error cases | 0.1 |
| $a_d$ | additive error deaths | $2 \times 10^{-4}$ |
| $b_d$ | multiplicative error deaths | 0.0754 |
| $a_{Ha}$ | additive error hospital admissions | $3.62 \times 10^{-3}$ |
| $b_{Ha}$ | multiplicative hospital admissions | 0.05 |
| $b_{Ho}$ | multiplicative hospital occupancy | 0.139 |

Table S3: Measurement error parameters for data creation with the SEIRAHD model. Note that hospital occupancy was only modelled with a multiplicative error, no additive error.[8] All error parameters are given on a normalized scale, i.e. they are applied to observations that have been scaled to 10000 population.

Thus, $\sigma^2(GI) = 26.01 + 25.5 + 25 = 76.51$ and $\sigma(GI) = 8.75$.

Since individuals in the hospitalized compartment are assumed not to be infectious, the contribution of the hospitalized compartment to the generation interval is 0. Thus, when using hospitalizations as observations, we used the same generation interval as for case observations, but lagged the NPIs by the average time from infection to hospitalization (10 days).

##### 1.4 Bootstrapping 2-step regression

To bootstrap, the 2-step regression approach, we first ran the $\mathcal{R}_t$ estimation normally. Next, ran 500 bootstrap iterations as follows: First, we sampled a constant quantile value from a Beta(2,2) distribution. Then, we extracted the quantile value of the $\mathcal{R}_t$ distribution ($\mathcal{R}_t$ is assumed to be gamma-distributed) for each weekly data point (illustrated in Figure S3A), and these $\mathcal{R}_t$ values were then used in the mixed effects model to estimate the NPI parameters. After 500 iterations, we calculated the 2.5th and 97.5th percentiles of the estimated parameters to derive lower and upper bounds of the CIs, respectively (Figure S3B).

##### 1.5 Next generation matrix approach

The next-generation matrix is a method to derive the basic or effective reproduction number for a compartmental model. For its calculation, only the "infected" compartments are used, so E, I, and A. Let $x_i, i = 1, 2, 3, ..., m$ be the numbers of infected individuals in the $i^th$ infected compartment at time t. Then, two matrices can be built: 1) $V_i(x)$, which represents the arrivals and departures from one of the infected compartments to another, and 2) $F_i(x)$, which describes the arrivals of new infections in compartment i. The matrices $V_i(x)$ and $F_i(x)$ are therefore

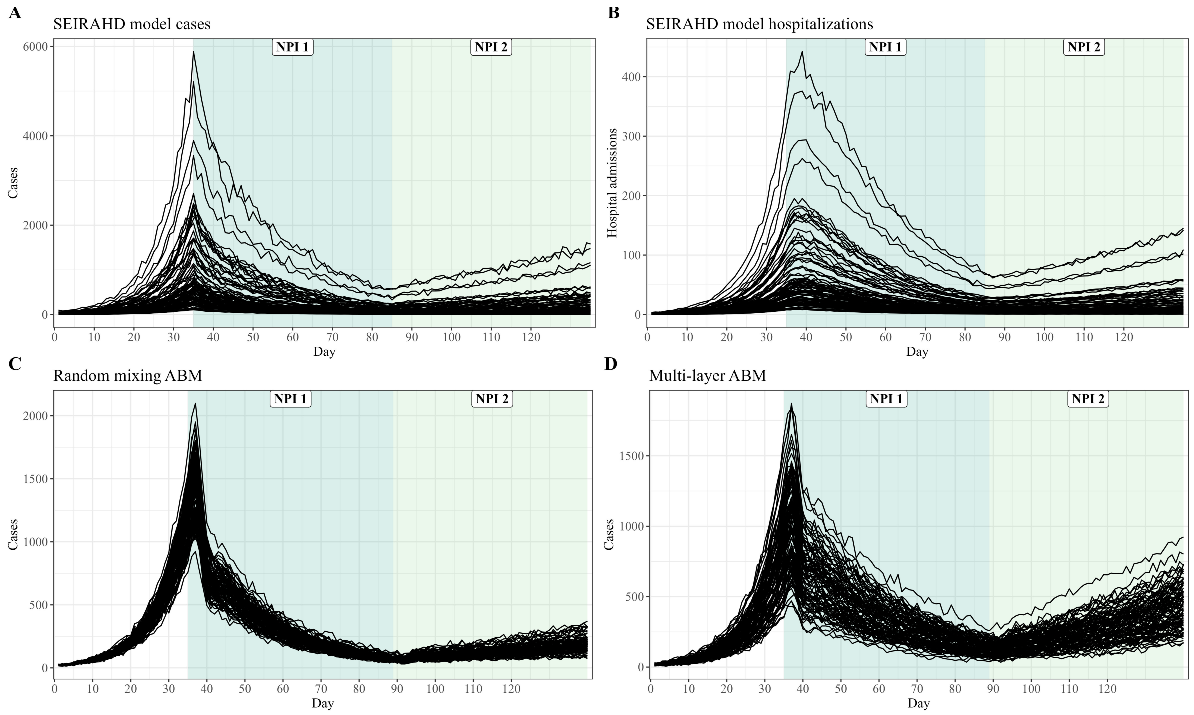


Figure S2: Example of created datasets with SEIRAHD model and ABMs. (A) Simulated cases with SEIRAHD model, (B) simulated hospitalizations with SEIRAHD model, (C) simulated cases with random mixing agent-based model, (D) simulated cases with layered agent-based model.

constructed as:[32]

$$V_i(x) = \begin{pmatrix} \frac{1}{D_E} & 0 & 0 \\ -\frac{r_E}{D_E} & \frac{r_H(1-e_{vH})}{D_Q} + \frac{(1-r_H)}{D_I} & 0 \\ -\frac{(1-r_E)}{D_E} & 0 & \frac{1}{D_I} \end{pmatrix} \tag{4}$$

$$F_i(x) = \begin{pmatrix} 0 & b\frac{S(1-e_{vI})}{N} & b\frac{\alpha S(1-e_{vI})}{N} \\ 0 & 0 & 0 \\ 0 & 0 & 0 \end{pmatrix} \tag{5}$$

Then, it has been shown that $\mathcal{R}_t = \rho FV^{-1}$, where $\rho FV^{-1}$ is the spectral radius (or largest eigenvalue) of the Next Generation Matrix $FV^{-1}$. One can picture the entries of $FV^{-1}$ as the rate at which infected individuals in $x_j$ produce new infections in $x_i$, times the average length of time an individual spends in compartment j. For a proof, see for example Perasso.[40] Therefore, we obtain:

$$\mathcal{R}_t = transmission(1-e_{vI})S(t)\left(D_I\alpha(1-r_E) + \frac{D_I D_Q r_E}{D_Q(1-r_H) + D_I(1-e_{vH})r_H}\right) \tag{6}$$

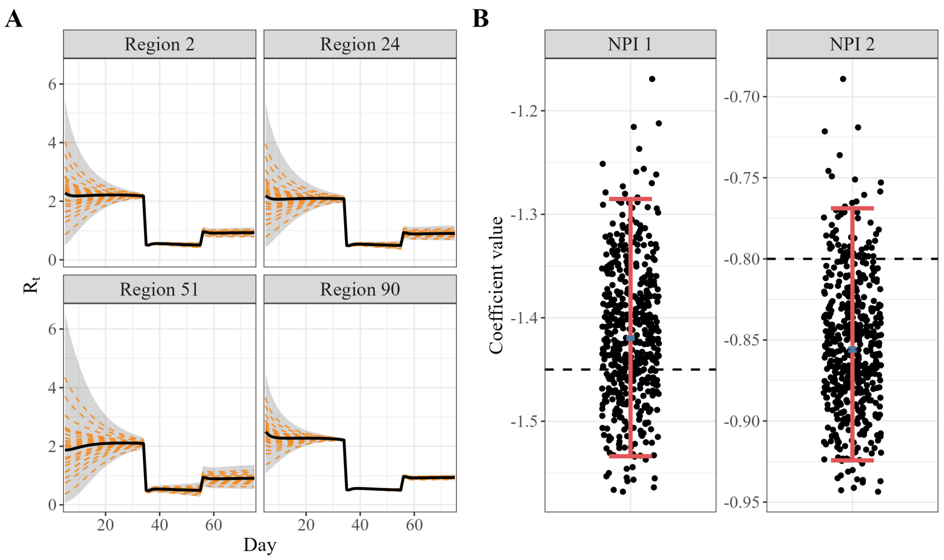


Figure S3: Illustration of the quantile bootstrap method. A: The $\mathcal{R}_t$ point estimate is shown as a solid black line, while the estimated 95% CI is depicted with the grey shaded area. Each dashed orange line represents the bootstrap draw of one constant quantile over time (only 50 iterations are shown for clarity). B: The NPI parameter distribution of all 500 bootstrap iterations is shown as black dots. The blue dot illustrates the point estimate and the red bars the 95% CI estimated from the bootstrap.

### 2 Supplementary Results

#### 2.1 SIR-generated data

#### 2.2 SEIRAHD-generated data

##### 2.2.1 Analysis with true $\mathcal{R}_t$

When the regression model was applied to ABM-created datasets, the bias in NPI effect estimation was larger, likely due to higher fluctuations in $\mathcal{R}_t$ (Figure S7).

##### 2.2.2 NPI implementation scenarios

##### 2.2.3 Incident infections vs. incident cases

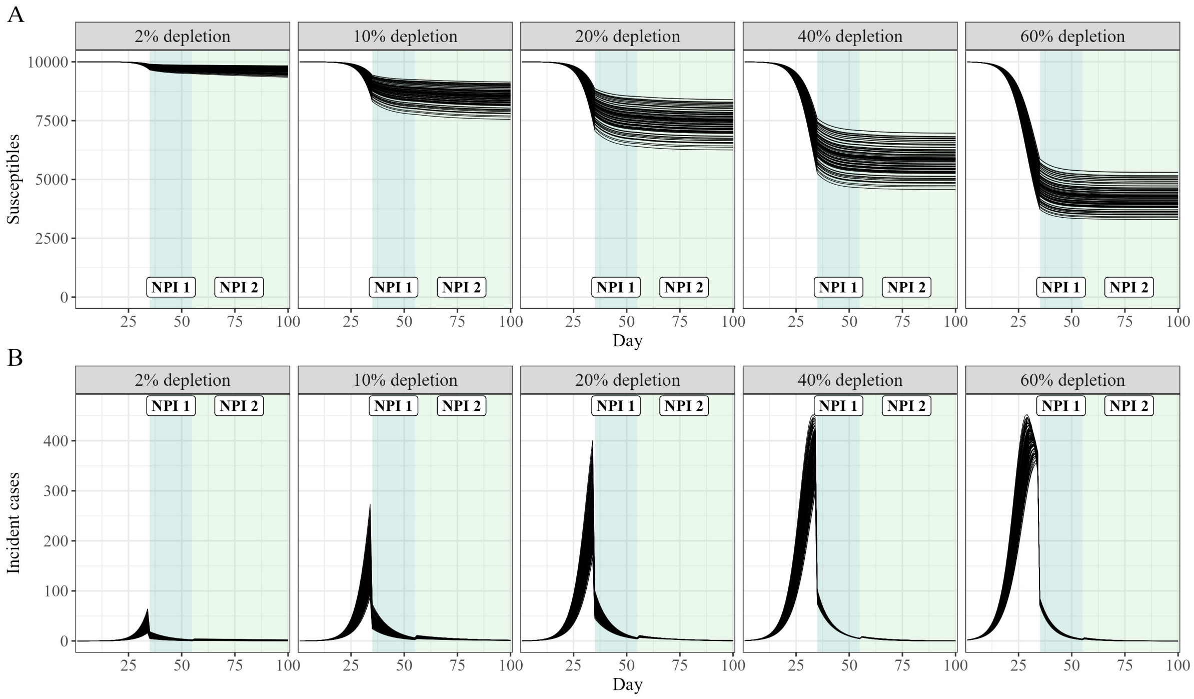


Figure S4: Example of created datasets with SIR models. (A) Plots of depletion of susceptibles, separated by depletion scenario, normalized to 10,000 population. (B) Plots of daily incident cases, separated by depletion scenario, with populations equally normalized to 10,000.

| Metric | SEIRAHD model | Random mixing ABM | Multi-layer ABM |
|---|---|---|---|
| **NPI 1** | | | |
| Absolute bias | 0.04 | 0.05 | -0.18 |
| Relative bias (%) | 2.9 | 3.6 | 12.3 |
| 95% CI (%) | 0 | 0 | 0 |
| **NPI 2** | | | |
| Absolute bias | 0.05 | 0.06 | -0.02 |
| Relative bias (%) | 6.7 | 6.9 | 2.9 |
| 95% CI (%) | 0 | 0 | 0 |

Table S4: Evaluation metrics for regressions with known $\mathcal{R}_t$ (average of 100 datasets). Note that the main part of the manuscript only describes results from SEIRAHD-created data, but ABM-generated data results were added for completeness.
ABM agent-based model, CI confidence interval, NPI non-pharmaceutical intervention

| | 1 week | | 2 weeks | |
|---|---|---|---|---|
| Metric | Gradient early | Gradient late | Gradient early | Gradient late |
| **NPI 1** | | | | |
| Absolute bias | -0.08 | -0.06 | -0.04 | -0.01 |
| Relative bias (%) | 5.6 | 4.4 | 3.0 | 0.6 |
| 95% CI (%) | 0 | 0 | 0 | 88 |
| **NPI 2** | | | | |
| Absolute bias | 0.03 | 0.10 | 0.06 | 0.15 |
| Relative bias (%) | 4.2 | 12.1 | 7.1 | 19.0 |
| 95% CI (%) | 26 | 0 | 0 | 0 |

Table S5: Results from 100 datasets with gradual NPI implementation, analyzed with the two-step regression method. NPIs were either implemented "early" (i.e. as in the main analysis after on day 16) or "late" (i.e. on day 27) and with a linear gradient either over 1 or 2 weeks.

| Metric | Estimation with incident cases | Estimation with incident infections |
|---|---|---|
| **NPI 1** | | |
| Absolute bias | -0.26 | 0.07 |
| Relative bias (%) | 18.3 | 4.5 |
| **NPI 2** | | |
| Absolute bias | -0.11 | 0.18 |
| Relative bias (%) | 13.7 | 22.9 |

Table S6: Comparison of bias in regression parameters, using incident cases (= entry into the I compartment) and incident infections (= entry into the E compartment)

[27] Cori, A. EpiEstim: Estimate Time Varying Reproduction Numbers from Epidemic Curves. 2021.

[33] Sam Abbott; Joel Hellewell; Katharine Sherratt; Katelyn Gostic; Joe Hickson; Hamada S. Badr; Michael DeWitt; Robin Thompson; EpiForecasts; Sebastian Funk EpiNow2: Estimate Real-Time Case Counts and Time-Varying Epidemiological Parameters. 2020.

```
File: main.tex
Encoding: utf8
Sum count: 6348
Words in text: 5007
Words in headers: 129
```

```
Words outside text (captions, etc.): 1081
Number of headers: 37
Number of floats/tables/figures: 20
Number of math inlines: 119
Number of math displayed: 12
Subcounts:
  text+headers+captions (#headers/#floats/#inlines/#displayed)
  205+20+0 (2/0/2/0) _top_
  404+1+0 (1/0/3/0) Section: Introduction} \label{introduction
  0+1+0 (1/0/0/0) Section: Methods} \label{methods
  564+19+0 (4/0/5/3) Subsection: Study design
  96+5+0 (1/0/2/1) Subsection: Parameter estimation with mechanistic models
  298+5+0 (1/0/10/1) Subsection: Parameter estimation with two-step regression
  42+2+0 (1/0/2/0) Subsection: Performance evaluation
  120+1+0 (1/0/1/0) Subsection: Implementation
  136+2+0 (1/0/4/0) Subsection: Bias exploration
  0+1+0 (1/0/0/0) Section: Results} \label{results
  526+17+132 (3/2/7/1) Subsection: Exploring bias in the two-step regression models
  421+3+124 (1/1/17/0) Subsection: Origins of bias
  190+11+49 (1/1/0/0) Subsection: Limitations of the mechanistic approach in the context of
  1036+1+0 (1/0/9/0) Section: Discussion} \label{discussion
  23+1+0 (1/0/0/0) Section: Contributions
  6+3+0 (1/0/0/0) Section: Declaration of interests
  15+2+0 (1/0/0/0) Section: Data sharing
  118+1+0 (1/0/0/0) Section: Acknowledgements
  0+1+0 (1/0/0/0) Part: Appendix
  0+2+0 (1/0/0/0) Section: Supplementary Methods
  0+2+5 (1/1/0/0) Subsection: SIR models
  210+2+94 (1/4/4/0) Subsection: SEIRAHD model} \label{appendix-SEIRAHD
  330+2+0 (1/0/24/3) Subsection: Generation intervals} \label{Generation intervals
  97+3+83 (1/1/5/0) Subsection: Bootstrapping 2-step regression} \label{appendix-bootstrap
  145+4+0 (1/0/12/3) Subsection: Next generation matrix approach}\label{NGM
```

```
  0+2+0 (1/0/0/0) Section: Supplementary Results
  0+2+151 (1/3/1/0) Subsection: SIR-generated data
  25+13+443 (4/7/11/0) Subsection: SEIRAHD-generated data

File: output.bbl
Encoding: utf8
Sum count: 707
Words in text: 707
Words in headers: 0
Words outside text (captions, etc.): 0
Number of headers: 0
Number of floats/tables/figures: 0
Number of math inlines: 0
Number of math displayed: 0

Total
Sum count: 7055
Words in text: 5714
Words in headers: 129
Words outside text (captions, etc.): 1081
Number of headers: 37
Number of floats/tables/figures: 20
Number of math inlines: 119
Number of math displayed: 12
Files: 2
Subcounts:
  text+headers+captions (#headers/#floats/#inlines/#displayed)
  5007+129+1081 (37/20/119/12) File(s) total: main.tex
  707+0+0 (0/0/0/0) File(s) total: output.bbl
```

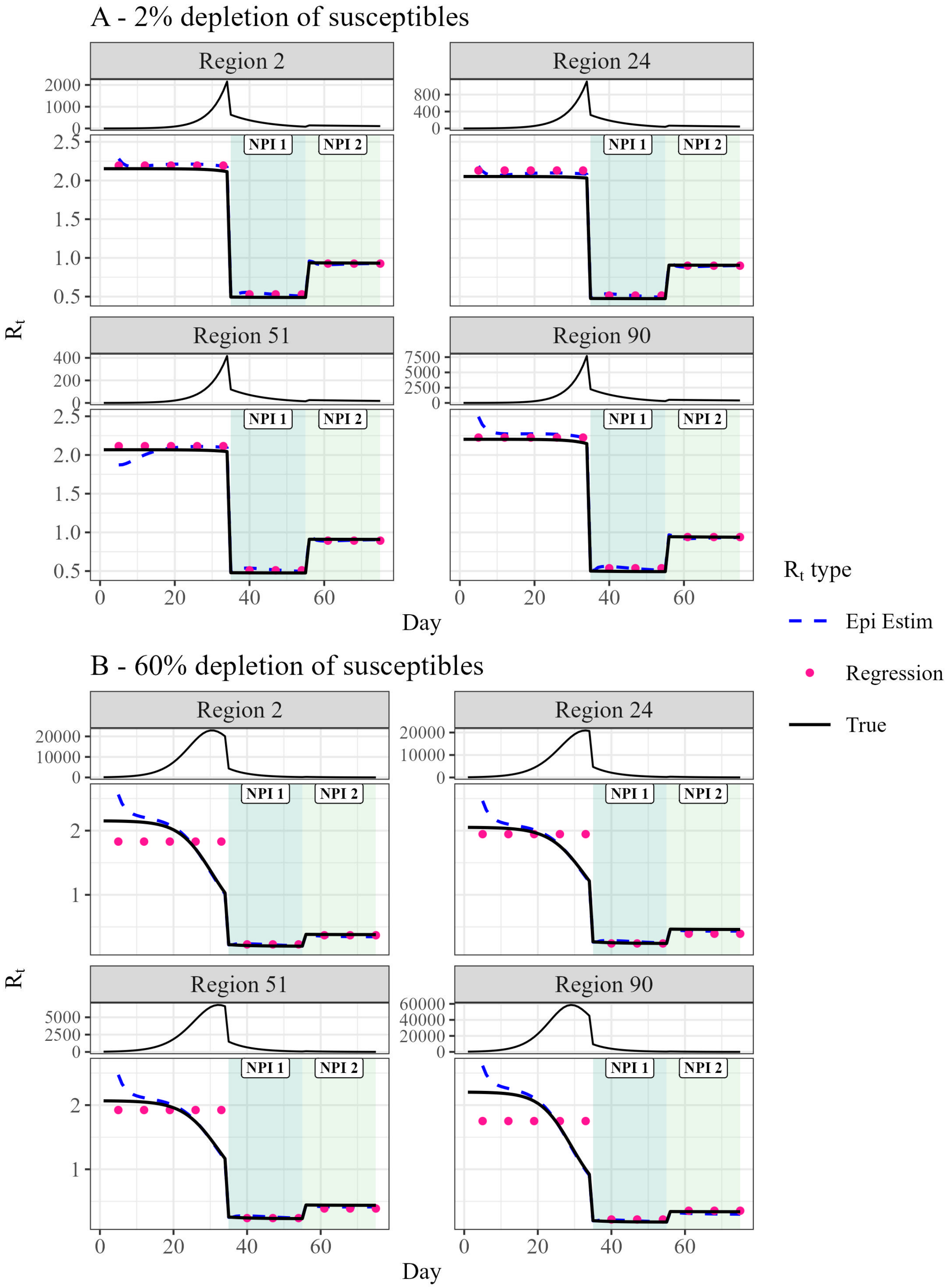


Figure S5: Comparison of $\mathcal{R}_t$ and regression fits from the two-step model from SIR-simulated datasets. A: Datasets generated with approx. 2% of susceptibles depleted at the time of NPI 1 implementation. B: Datasets generated with approx. 60% of susceptibles depleted at the time of NPI 1 implementation. Each panel represents one geographic region. The highlighted regions indicate which NPI was active at which time. The panels on top show the respective case time series. NPI: non-pharmaceutical intervention

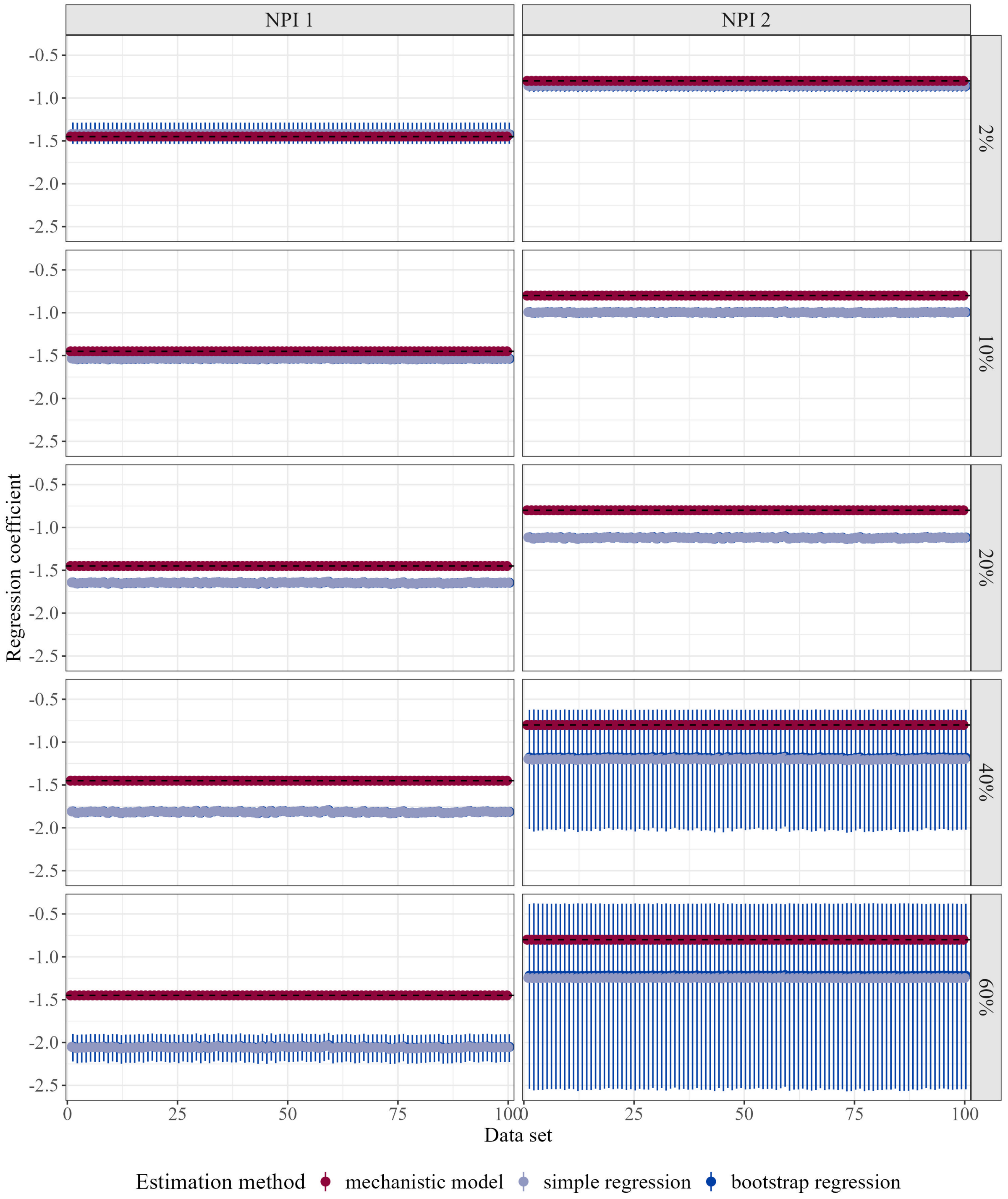


Figure S6: Estimation results from SIR-generated data under different scenarios of depletion of susceptibles. For each NPI separately, the point estimate and 95% CIs are shown for each estimation method. The dashed black lines indicate the value used in simulation.

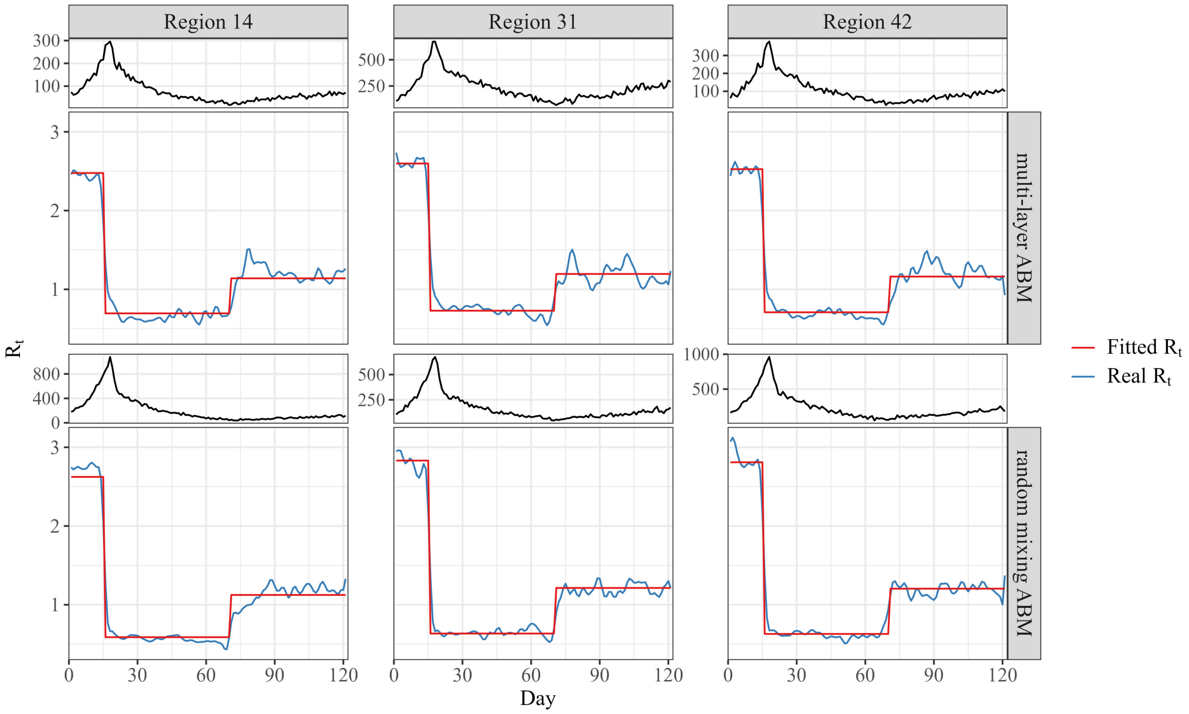


Figure S7: Regression fits of true $\mathcal{R}_t$ in three randomly selected regions. Each panel represents one geographic region with data generated by either a random mixing ABM or a multi-layer ABM. The true $\mathcal{R}_t$ is depicted in blue and the corresponding regression fit in red. The panels on top show the respective case time series.

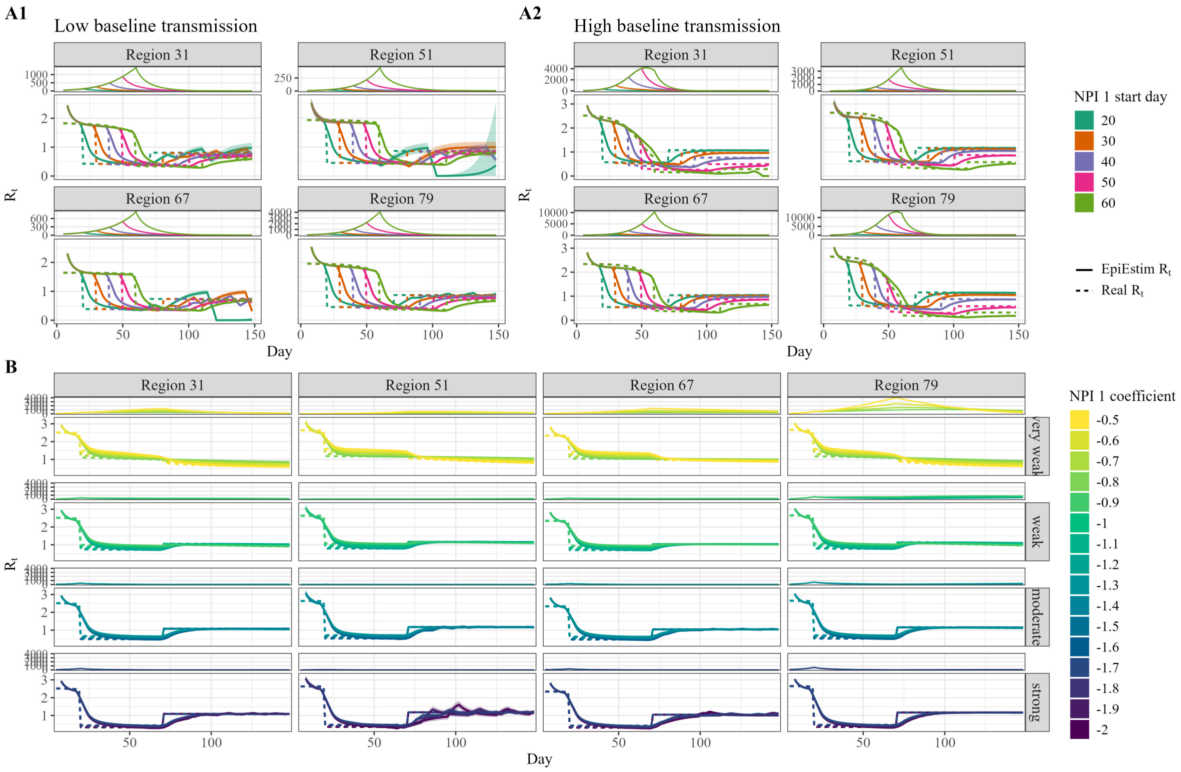


Figure S8: $\mathcal{R}_t$ fits by EpiEstim for varying NPI 1 scenarios. Scenarios are shown for four selected geographical regions, indicated by the numbers above the panels. The panels on top depict the case time series. **A1:** Scenarios are shown for varying NPI 1 start days where the basic transmission rate was adjusted to prevent a reduction in $\mathcal{R}_t$ due to high population immunity. **A2:** Scenarios for varying NPI 1 start days, with the basic transmission rate remaining consistent with the main analysis. Especially in the later NPI implementation scenarios, a notable decrease in $\mathcal{R}_t$ by population herd immunity becomes evident. **B:** $\mathcal{R}_t$ estimates for three selected regions in scenarios with varying NPI 1 strength. Considering the presentation of numerous scenarios, we organized the trajectories into four rows based on NPI strength.
NPI non-pharmaceutical intervention

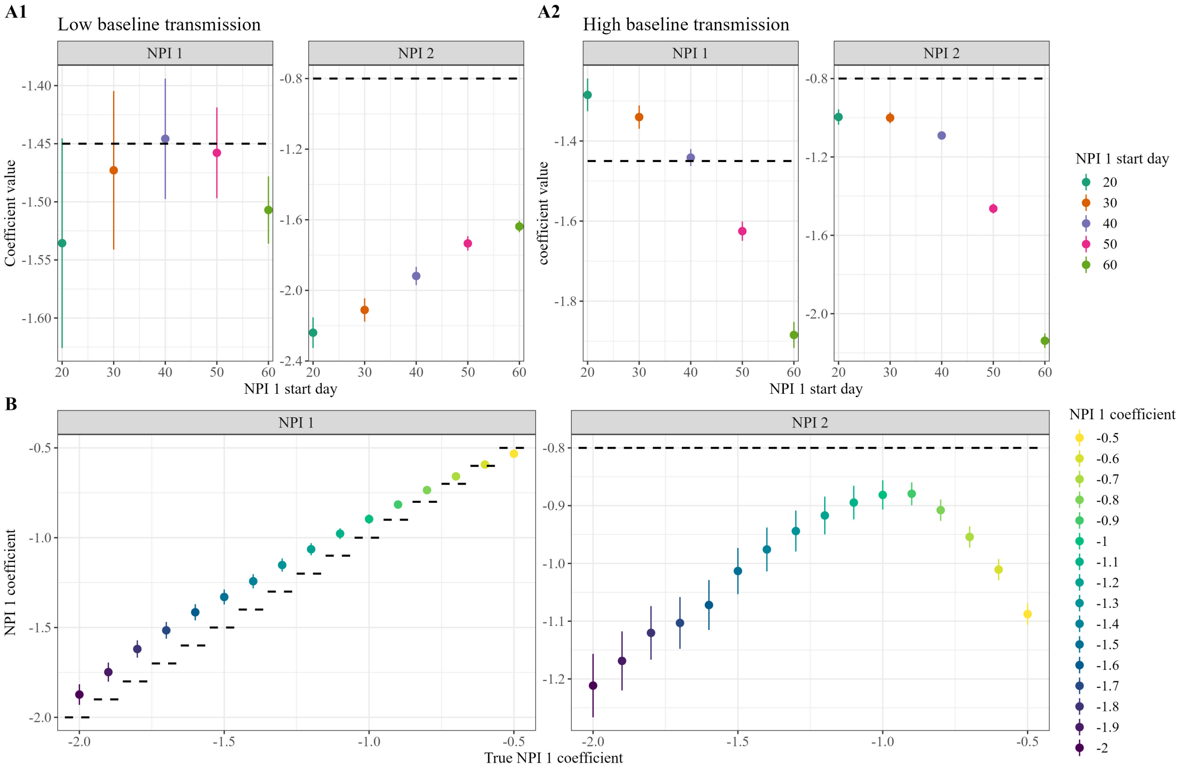


Figure S9: Regression coefficients from the two-step regression for varying NPI 1 scenarios. Results are shown separately by NPI. The dashed lines indicate the true NPI values. **A1:** Coefficients with corresponding 95% CIs are shown for varying NPI 1 start days where the basic transmission rate was adjusted to prevent a reduction in $\mathcal{R}_t$ due to high population immunity. **A2:** Coefficients with corresponding 95% CIs for varying NPI 1 start days, with the basic transmission rate remaining consistent with the main analysis. **B:** Coefficients with corresponding 95% CIs for scenarios with varying NPI 1 strength. Considering the presentation of numerous scenarios, we organized the trajectories into four rows based on NPI strength.
NPI non-pharmaceutical intervention

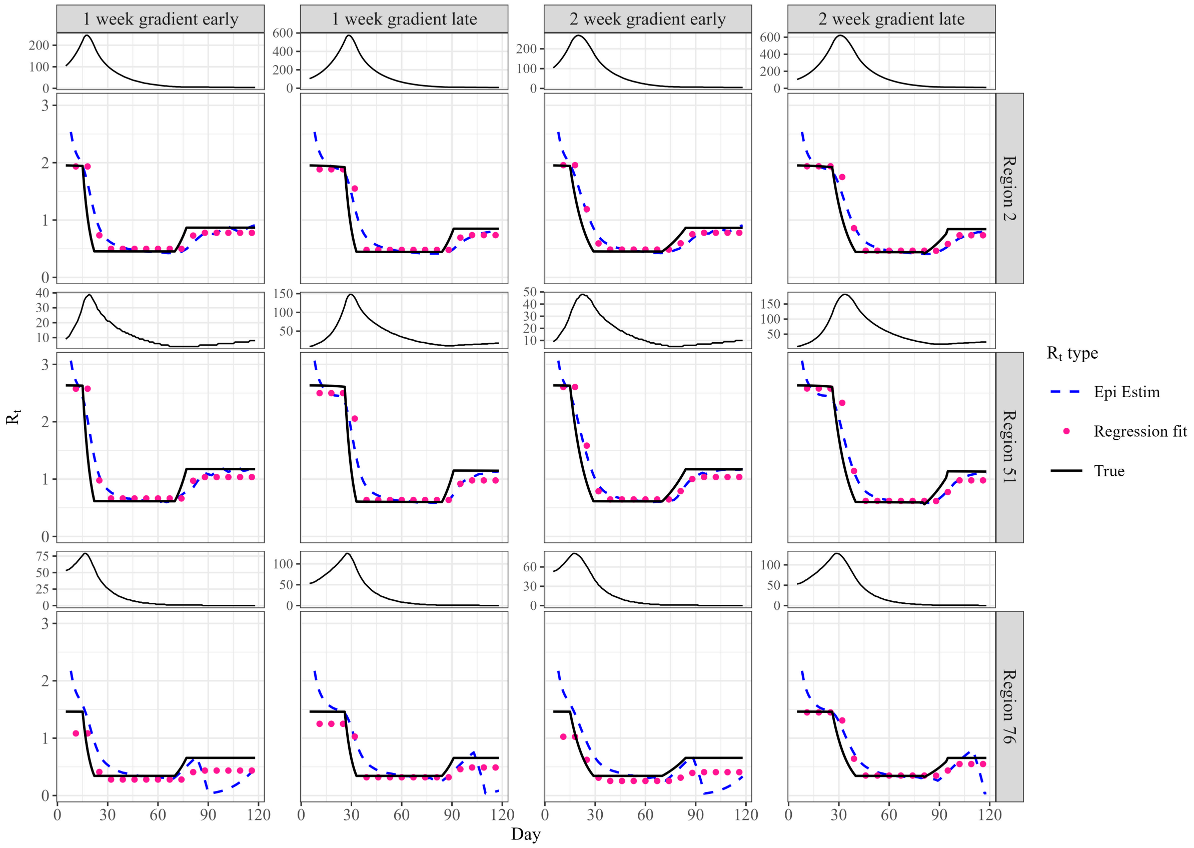


Figure S10: $\mathcal{R}_t$ and regression fits from the two-step model with gradual NPI implementation. NPIs were either implemented ”early” (i.e. as in the main analysis after on day 16) or ”late” (i.e. on day 27) and with a linear gradient either over 1 or 2 weeks.
